# Dry Season Mosquito Breeding Ecology in the Sunyani Municipality, Bono Region, Ghana

**DOI:** 10.64898/2025.12.11.25342015

**Authors:** Samuel Yaw Agyemang-Badu, Esi Awuah, Sampson Oduro-Kwarteng, Nazri Che Dom, Justice Yao Woelinam Dzamesi, Samuel Offei

**Affiliations:** College of Health-Yamfo. Department of Community Health. Ministry of Health, Health Training Institution, Sunyani-Yamfo. Ghana; Regional Water and Environmental Sanitation Centre (RWESCK), World Bank African Centre of Excellence (ACE), Kwame Nkrumah University of Science and Technology (KNUST), Kumasi, Ghana; Faculty of Health Sciences, Universiti Teknologi MARA, UITM Cawangan Selangor, Malaysia; Environmental Health and Sanitation Unit, Tano South Municipal Assembly (TSMA), Bechem, Ahafo Region, Ghana

**Keywords:** Mosquitoes, Breeding Sites, Vector-Borne Diseases, Anthropogenic, Sunyani Municipality, Larval Source Management, Ghana

## Abstract

Mosquito-borne diseases (MBDs) remain a significant public health burden worldwide. This study aimed to survey mosquito breeding sites and assess larval distribution within the Sunyani Municipality, Bono Region, Ghana, to understand their potential contribution to disease transmission.

A survey of mosquito breeding sites and larval sampling was conducted using a simple random sampling technique during the dry season (December 2019 to February 2020). Daily inspections of habitats were carried out between 8:00 a.m. and 5:00 p.m. using the dipping method for larval collection. Mosquito breeding sites were identified based on standard guidelines, while larvae were morphologically identified using taxonomic keys.

*Anopheles* larvae were predominantly found in natural wetlands (64%), followed by natural drains with partially clean water (19%), and burst or leaking water distribution pipelines (17%). Of the total positive mosquito breeding sites, 31%, 58%, and 11% were attributed to *Anopheles, Culex*, and *Aedes*, respectively. From the 1,555 larvae sampled, *Culex* accounted for the majority (53%), followed by *Anopheles* (32%) and *Aedes* (15%). The distribution of larvae across breeding sites indicates a potential impact on the prevalence of vector-borne diseases, particularly malaria and yellow fever, in the Municipality.

During the dry season, mosquito breeding sites were primarily natural for *Anopheles* and anthropogenic for *Culex* and *Aedes*, highlighting the influence of environmental mismanagement and poor enforcement of sanitation regulations. There is an urgent need for extensive community education on the health risks associated with mosquito breeding sites. Integrated mosquito control measures, including larval source management, removal of breeding habitats, and larvicide application, should be prioritized as part of a comprehensive elimination program in the Municipality.

## Introduction

Mosquito-borne diseases (MBDs), including malaria, dengue, yellow fever, and Zika virus, remain significant public health challenges worldwide, particularly in tropical regions where environmental conditions favor mosquito proliferation ^[1,2]^. Malaria, primarily transmitted by female *Anopheles* mosquitoes, continues to impose a substantial burden in sub-Saharan Africa, while *Aedes aegypti* and *Aedes albopictus* are major vectors of arboviral diseases such as dengue and Zika virus, exacerbating health crises in densely populated areas ^[3]^. These diseases are linked to high morbidity and mortality rates, economic losses, and strain on healthcare systems, highlighting the urgent need for effective vector control strategies ^[4,5]^. Globally, vector control has relied on approaches such as larval source management, adult mosquito control through insecticide spraying, and community sanitation efforts. However, these methods face limitations, including the emergence of insecticide resistance, changes in breeding habitats due to rapid urbanization, and gaps in entomological surveillance, particularly at the local level ^[6,7]^. In Ghana, recent outbreaks of yellow fever and the endemicity of malaria underscore the pressing need for localized solutions tailored to address vector proliferation and disease transmission dynamics ^[8]^.

The Sunyani Municipality, located in the Bono Region of Ghana, presents a unique challenge due to its environmental and anthropogenic factors, which provide an abundance of mosquito breeding sites. Despite the known association between these breeding habitats and disease transmission, studies characterizing larval ecology and identifying species composition at the local level remain scarce ^[9]^. Understanding these ecological features is crucial for designing targeted and sustainable mosquito control programs. This study aims to address these gaps by virtually sampling mosquito larval habitats and morphologically identifying mosquito species of medical importance within the Sunyani Municipality. By assessing population density, dynamics, and prevalence, the study seeks to provide empirical data to inform vector control strategies, ultimately contributing to the reduction of MBDs in the region. Insights gained from this study are expected to support the development of targeted and sustainable vector control strategies aimed at mitigating the high disease burden in the region.

## Materials and Methods

### Study Area

The study was conducted in the Sunyani Municipality, located in the Bono Region of Ghana. Geographically, the municipality lies between latitudes 7°20’N and 7°05’N and longitudes 2°30’W and 2°10’W, covering a total area of 506.7 km². Sunyani serves as the regional capital and is predominantly urban, with over 80% of the population residing in urbanized areas. This urban dominance facilitates dynamic interactions between human activities and the surrounding environment, which contribute to the creation of ideal mosquito breeding habitats. The Municipality is subdivided into multiple zonal councils and communities, such as Newtown, Estate, Baakoniaba, Atronie, and Penkwase, which are characterized by diverse land-use patterns, including residential, commercial, and industrial zones (**Fig. 1**) ^25-26]^.

**Fig. 1:**
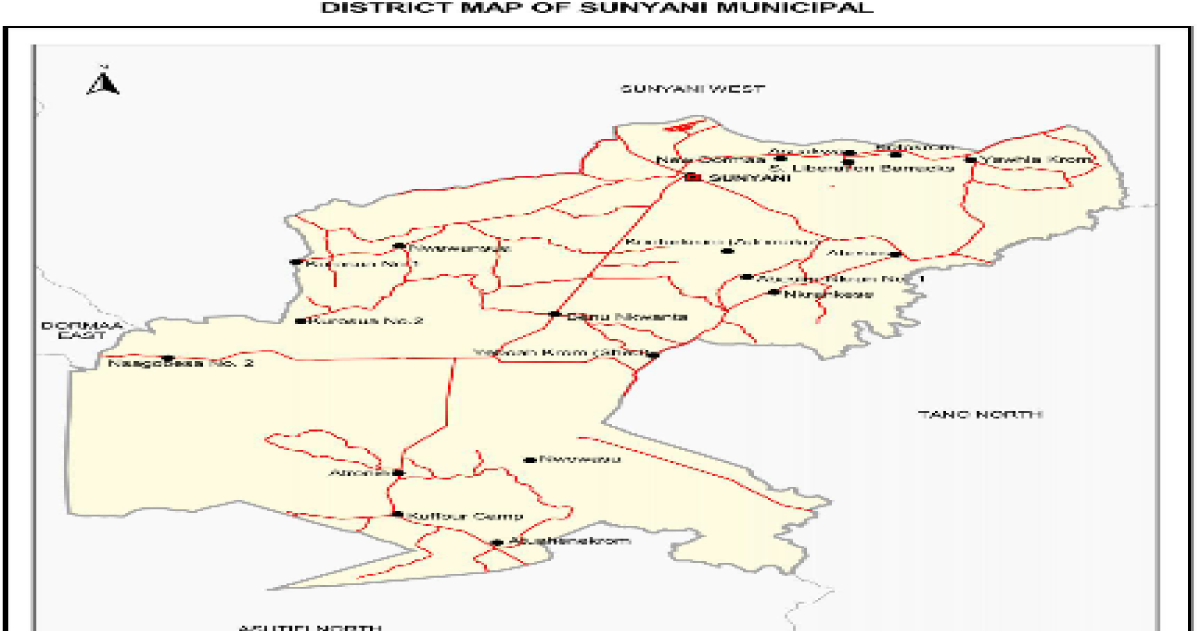
Map of Sunyani Municipality ^[25–26]^.

The region experiences a tropical climate, with distinct wet and dry seasons. Rainfall follows a double maxima pattern, with the main rainy season occurring from March to September and a minor season extending from October to December, averaging 88.99 cm annually. Temperatures range between 23°C in August and 33°C in March and April, with relative humidity fluctuating between 75% and 80% during the rainy seasons and dropping below 70% in the dry seasons. These climatic conditions, coupled with the municipality’s location in the moist semi-deciduous forest zone, provide an optimal environment for mosquito breeding. Natural features such as swampy areas, waterlogged lands, and lush vegetative growth enhance the availability of breeding habitats, while anthropogenic activities exacerbate mosquito proliferation ^[25–26].^

The population of Sunyani Municipality, based on recent census data, stands at 123,224, with an almost equal distribution of men (61,610) and women (61,614) ^[25–26]^. The region’s socioeconomic activities, including agriculture, trade, and construction, inadvertently contribute to the prevalence of mosquito breeding sites. Improper disposal of water containers, poor drainage systems, and stagnant water around construction areas create artificial habitats conducive to mosquito larval development. Additionally, the widespread use of water storage containers during dry periods further amplifies breeding opportunities. Sunyani Municipality was selected for this study due to its significant incidence of mosquito-borne diseases, as reported by the Bono Regional Health Directorate. The area’s mix of natural and anthropogenic breeding habitats provides an ideal setting for studying mosquito ecology and population dynamics ^[25–26]^.

### Sampling Methodology

A systematic sampling technique was employed to investigate mosquito breeding habitats, population density, and genus (species) diversity within the three zonal councils, made up of twenty-one (21) communities across the Sunyani Municipality, where the researchers were moving sluggishly and judiciously in the communities to ascertain the presence or absence of immature mosquito larvae, by identifying, inspecting and examining water bodies, recipients (containers/ receptacles), wetlands (swampy and marshy areas), drains and water supply and storage facilities with the potential to harbor mosquito larvae. Sampling was conducted between December 2019 and February 2020 in the communities categorized into five zones (**Table 1**). This period was strategically chosen as it coincided with the dry season, a time when water bodies and artificial containers often become critical breeding sites for mosquitoes. The communities sampled included urban and semi-urban areas such as Newtown, Estate, Baakoniaba, Penkwase, and Abesim, ensuring a representative assessment of diverse environmental and anthropogenic settings. However, the land assessed included private owned and public areas (lands), and no protected land nor species were sampled during the study. Further, animal husbandry, experimentation, and care/welfare, were not relevant in this study.

**Table 1.** Distribution of Communities by Zone Sampled Across Sunyani Municipality.

| **Zones** | **Communities Covered** |
| --- | --- |
| Zone 1 | Newtown |
|  | Estate |
|  | Baakoniaba |
|  | Atronie |
|  | Asufufu |
| Zone 2 | Area 1 |
|  | Area 2 |
|  | Area 3 |
|  | Area 4 |
|  | Magazine |
|  | Wednesday Market |
| Zone 3 | Zongo Abetifi |
|  | Nkwabeng North |
|  | Nkwabeng South |
|  | Berlin Top |
| Zone 4 | Penkwase |
|  | New Dormaa |
|  | Asuakwa |
|  | Kotokrom |
|  | Yawhima |
| Zone 5 | Abesim |
| **Total** | **21** |

**Note:** The table highlights the number of communities per each zonal area sampled during the study period (December 2019 to February 2020).

In this study, mosquito breeding habitat/site, is a larval habitat, where developmental stages of mosquitoes (eggs, larvae, pupae) are found, including sites that appear to be ecologically suitable for varied species. Also, positive larval habitat, is a water body and/or recipient with mosquito larvae or pupae found alive during the field larval sampling, while negative/potential habitat/site is a habitat/site suitable of producing mosquito larvae or pupae, but larvae or pupae were not found. However, positive mosquito larval sample, is a live larvae or pupae collected through the dipping method, transported to the laboratory, and reared to the adult stage.

Field sampling involved systematically identifying and inspecting potential mosquito breeding habitats, including natural sites such as wetlands, marshy areas, and stagnant water bodies, as well as anthropogenic sites like water recipients, and leaking water pipes. Each site was visually inspected for the presence of mosquito larvae, which were collected using the dipping method ^[27, 28]^. A calibrated 10 cm diameter ladle with a 90 ml capacity was used to take 2–3 dips, with an average of 3 dips per site, along the edges of each breeding site, depending on its size. In all, a total of 4,122 dips were taken for both positive and negative breeding sites identified and sampled. The collected larvae were transferred into labelled 250 ml plastic containers for transport to the laboratory, with labels indicating the species group (e.g., *Anopheles* or *Culicinae)* and site of collection **(Fig. 2A)**. Sampling activities were conducted daily between 8:00 a.m. and 5:00 p.m., with each site inspected for approximately 20 minutes ^[27–28]^.

**Fig. 2:**
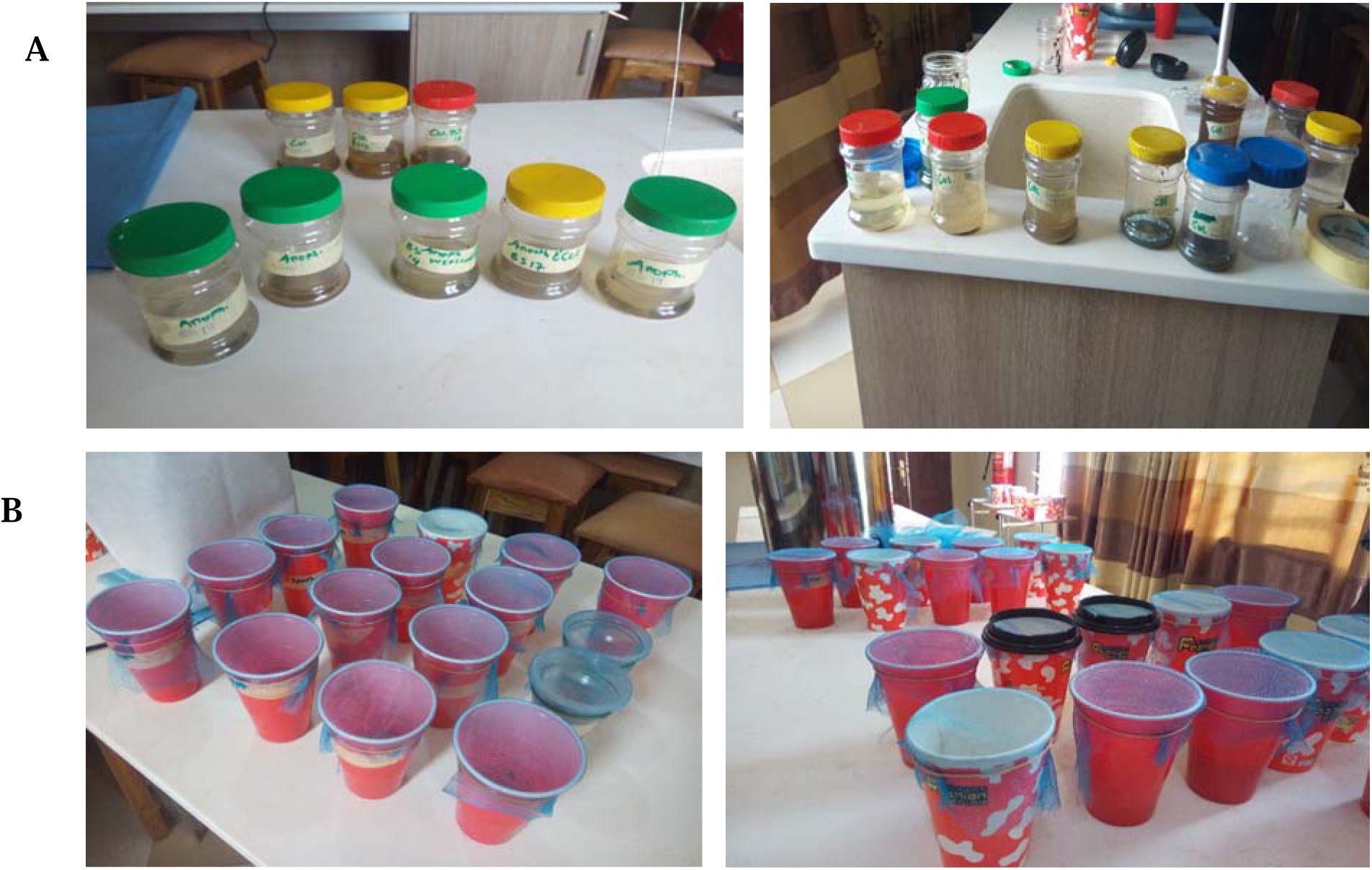
(A) Mosquito larval samples from the field at the laboratory ready for processing (rearing to adult stage) for all genus (species) and (B) Processing of mosquito larval samples at the laboratory (larva growing through pupation to adult stage) for morphological identification for all genus (species).

### Mapping Breeding Sites

To understand the spatial distribution of mosquito breeding sites within the Sunyani Municipality, a mapping exercise was conducted to identify positive and negative sites based on larval presence. Each breeding site was geo-referenced using the GPS Essential application, a reliable tool for capturing spatial coordinates. Data for positive breeding sites, including mosquito genus (*Anopheles*, *Culex*, or *Aedes*), were recorded in a field data logbook. GPS coordinates for each site, whether positive (larvae or pupae present) or negative (no larvae or pupae found), were captured using the GPS Essential mobile software application. GPS coordinates were subsequently exported to Google Earth to generate detailed maps illustrating the spatial distribution of breeding sites across the municipality. These maps highlighted high-risk zones and the relationship between breeding site locations and environmental features, such as water bodies, vegetation, and urban infrastructure.

At each identified breeding site, site-specific characteristics such as habitat type, water quality (e.g., clear or turbid), vegetation presence, and proximity to human dwellings were observed. Positive breeding sites were further classified based on the presence of larval species, and larvae were reared to adulthood for precise morphological identification **(Fig. 2)**. These classifications provided critical insights into the spatial clustering of mosquito habitats and their environmental determinants.

### Environmental Variable Assessment

Environmental variables were assessed to characterize mosquito breeding habitats and understand factors influencing larval development in the Sunyani Municipality. Each identified breeding site was evaluated based on its physical, biological, and environmental attributes. Breeding habitats were categorized as natural (e.g., wetlands, marshy areas, and rain pools) or anthropogenic, that is, water recipients (e.g., containers, discarded receptacles, drains, and storage facilities). Habitat permanence was visually classified as temporary or permanent based on the presence and persistence of standing water. Water depth were visually observed, and turbidity was regarded as either clear or turbid based on visual inspection. Light conditions at each site were visually examined as exposed to sunlight or shaded, depending on the presence of canopy cover or nearby structures ^[27–28]^.

Water quality parameters, including electrical conductivity, were observed to assess the suitability of the habitat for mosquito larvae. Additionally, the presence of foul odor were also observed and noted as an indicator of organic pollution. Biological components such as algae and aquatic plants were visually observed, with vegetation regarded as emergent, floating, or submerged. Sites lacking vegetation were also recorded. The presence of green algae, which serves as a food source for mosquito larvae, was visually observed and noted for each site. The surrounding environment was analysed by visually estimating the proximity of breeding sites to human dwellings. This parameter is critical as it correlates directly with the risk of human-vector interactions. Hydrological conditions were also assessed, with water current visually categorized as stagnant or slow-flowing, as stagnant water is typically associated with mosquito larval habitats ^[27–28]^.

All data were documented using a standardized checklist to ensure consistency. Variables recorded included site GPS coordinates, habitat type, location/area, number of larvae sampled, number of dips, type of mosquito genus found, etc. These parameters were analysed to identify environmental determinants influencing mosquito breeding site suitability ^[28]^. The comprehensive dataset provided insights into the ecological factors that support larval development, which are essential for developing targeted vector control strategies. These findings contribute to the understanding of mosquito ecology and inform sustainable interventions to reduce the prevalence of mosquito-borne diseases in the region.

### Mosquito Rearing and Genus Identification

The collected mosquito larvae were processed in the laboratory to ensure accurate identification and classification into genus of medical importance. Upon arrival from the field, larvae were transferred to labeled plastic recipients covered with a netting lid to prevent escape upon adult emergence, as shown in **Fig. 2B and 3**. Each recipient was filled with clean water to replicate natural aquatic environments, and a small plastic bottle was placed inside the recipient to provide a resting surface for emerging adults. The larvae were fed with a maize-based Cerelac solution to simulate the organic nutrients typically found in their natural habitats and ensure survival during rearing. However, mosquito larval densities (LD) were measured as the total number of larvae divided by the number of dips.

**Fig. 3:**
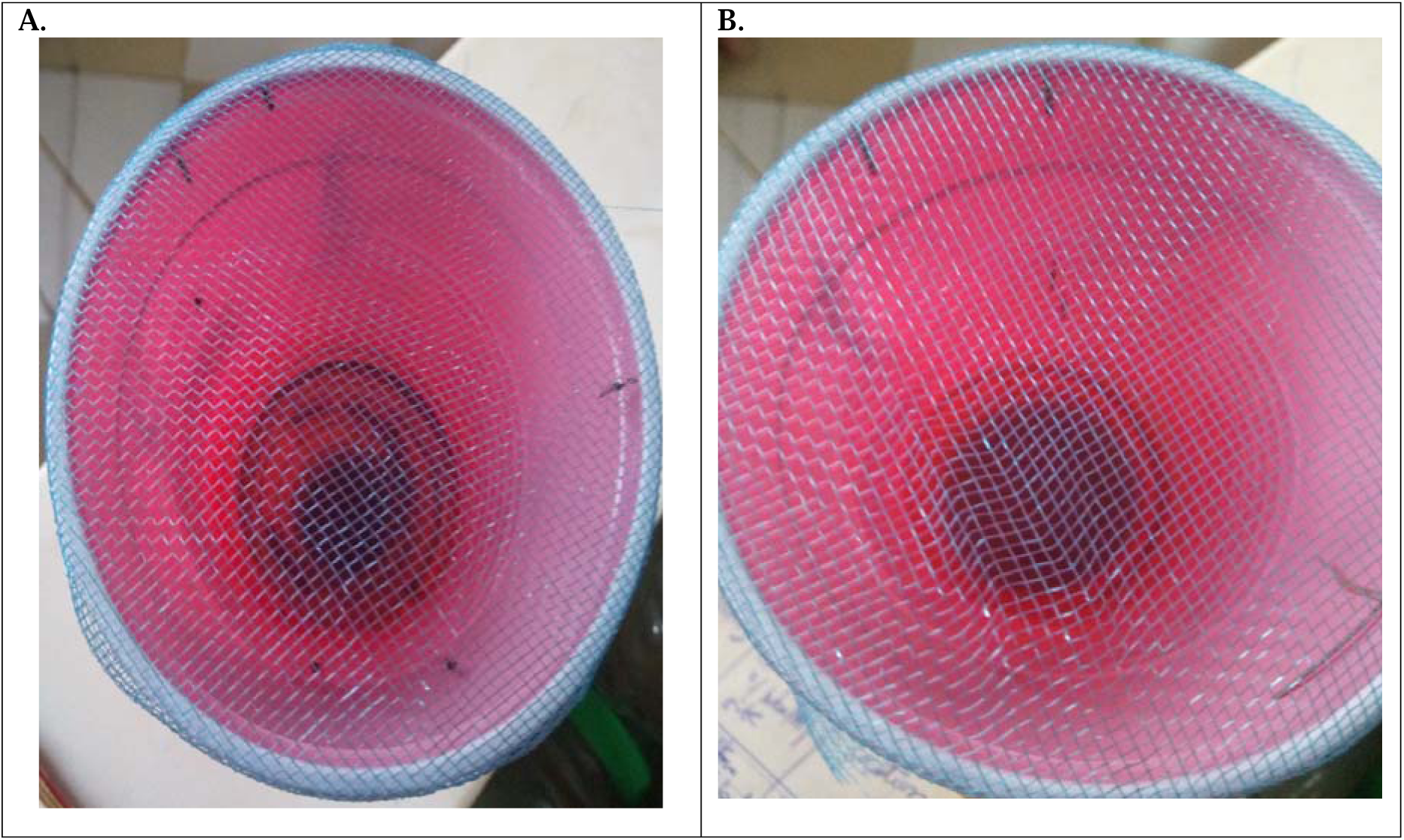
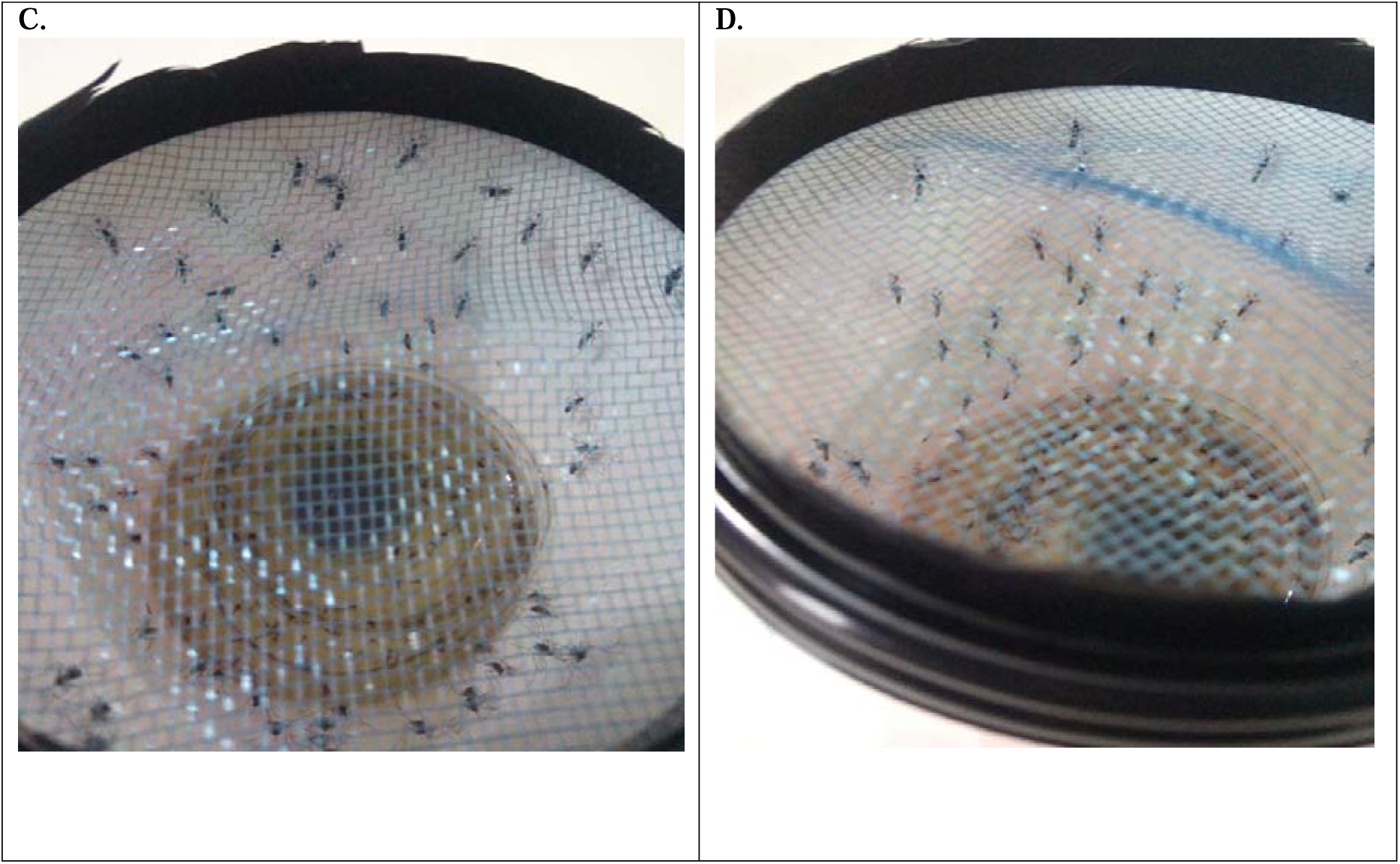
Fully grown adult mosquitoes in a plastic recipients covered with a net at the laboratory. (A), and (B) adult *Anopheles* mosquitoes resting at an angle to the resting surface. (C), and (D) adult *Culex* mosquitoes resting parallel to the resting surface.

The larvae were monitored daily, and those in advanced stages (instar III and IV) were observed for molting into pupae. Pupae, which represent a non-feeding stage, were carefully maintained until the adult mosquitoes emerged. The emergence process typically took 2–4 days for pupae, while early-stage larvae (instar I and II) required 7–14 days to complete development ^[27]^. Morphological characteristics such as body size, wing patterns, and leg markings were also examined under a stereomicroscope for precise genus level classification ^[30]^. The reared adults were allowed to dry and stabilize within the recipients, which were placed in an environment with controlled temperature and relative humidity of 27°C ± 2°C (80°F ± 2° F) and 76% ± 4% respectively, as shown in **Fig. 3**. Adult’s genus identification was conducted using standard morphological taxonomic keys by Knight and Stone ^[31–34]^.

Both *Anopheles*, *Culex*, and *Aedes* genus were identified and documented in accordance with standard taxonomic guidelines by ^[30]^, and validated against a pre-designed checklist. For habitats producing mixed genus, identification of each genus was meticulously recorded, along with the associated habitat type.

### Ethics Statements

The study was conducted in accordance with the guidelines of the Declaration of Helsinki and School of Graduate Studies, KNUST, and approved by the Committee on Human Research Publication and Ethics (CHRPE), School of Medical Sciences, Kwame Nkrumah University of Science and Technology (KNUST), Kumasi, Ghana with protocol code/reference number *CHRPE/AP/143/20.* However, there were no human participants involved in the study.

### Community Entry and Consent for Larval Sampling in both Private and Public Owned Areas

An official written notification letters were sent to the Regional Director of Health Services (RDHS), the Regional Environmental Health and Sanitation Officer (REHO), The Municipal Director of Health Services (MDHS) and the Municipal Environmental Health and Sanitation Officer (MEHO) for the Sunyani Municipality, where the aforementioned Officers offered verbal consent and their full support for the research work to be undertaken in the study area (s) which were under their various jurisdictions, since Ethical approval had already been granted by the School of Medical Sciences, KNUST as mentioned above.

### Limitation of the Study

While morphological identification was prioritized, molecular techniques such as rDNA Polymerase Chain Reaction (PCR) were not performed due to resource constraints, limiting identification to only genus levels based solely on morphological features ^[35–36].^ The detailed mosquito rearing (laboratory processing) and identification ensured reliable data on mosquito genus composition, supporting the ecological and spatial analysis of breeding habitats. These findings were instrumental in understanding mosquito population dynamics and informing targeted vector control strategies within the study area.

### Data Analysis

The collected data were systematically analysed to evaluate mosquito breeding habitat characteristics, larval abundance, species diversity, and spatial distribution within the Sunyani Municipality. Descriptive statistics were employed to summarize and present key findings, including the total number of breeding sites sampled, the proportion of positive and negative sites, and the distribution of mosquito species across different habitat types. Data on environmental variables such as water depth, turbidity, vegetation presence, and proximity to human dwellings were also analyzed to identify potential factors influencing larval presence.

To assess the spatial dynamics of mosquito breeding sites, geospatial analysis was performed using Google Earth and Geographic Information System (GIS) software. GPS coordinates of breeding sites were plotted to generate maps showing the distribution of positive and negative habitats. These maps were overlaid with environmental features such as urban structures, vegetation, and water bodies to provide visual insights into habitat suitability and clustering of breeding sites. The spatial analysis also highlighted high-risk areas, enabling targeted recommendations for vector control interventions. Comparative analysis was conducted to evaluate the relative abundance of different mosquito species (*Anopheles*, *Culex*, and *Aedes*) and their association with specific habitat characteristics. For example, genus (species)-specific preferences for shaded or sunlit habitats, natural or artificial breeding sites, and water quality parameters (e.g., turbidity, pH) were examined to identify ecological patterns. Correlations between environmental variables and mosquito presence were statistically tested to determine significant predictors of breeding site suitability.

The temporal dynamics of larval development and emergence were also analysed by monitoring the rearing process in the laboratory. This analysis provided additional insights into species-specific life cycle characteristics and their implications for vector control strategies. The integration of field and laboratory data ensured a comprehensive understanding of mosquito ecology and population dynamics. Overall, the data analysis provided actionable insights into the environmental and spatial factors driving mosquito breeding in the Sunyani Municipality. These findings are critical for designing evidence-based and localized vector control strategies to mitigate mosquito-borne diseases in the region.

## Results

### Distribution of Mosquito Breeding Sites Across Communities in the Sunyani Municipality

In all, a total of 1,374 potential (both positive and negative) mosquito breeding sites were identified and sampled across the municipality. In total, 555 positive mosquito breeding sites were identified and sampled, while on the other hand, 819 breeding sites sampled were found to be negative, across 21 communities in Sunyani Municipality, as detailed in **Table 2**. The distribution of these sites varied significantly across the municipality, reflecting the influence of environmental and anthropogenic factors. Among the communities, Penkwase recorded the highest number of positive breeding sites, with 73 sites accounting for 13.2% of the total. Following Penkwase, Nkwabeng North and Baakoniaba recorded 54 (9.7%) and 46 (8.3%) positive breeding sites, respectively, indicating these areas also serve as significant contributors to mosquito populations. Conversely, communities such as Area Three recorded the least number of positive breeding sites, with only 5 (0.9%) sites identified.

**Table 2.**
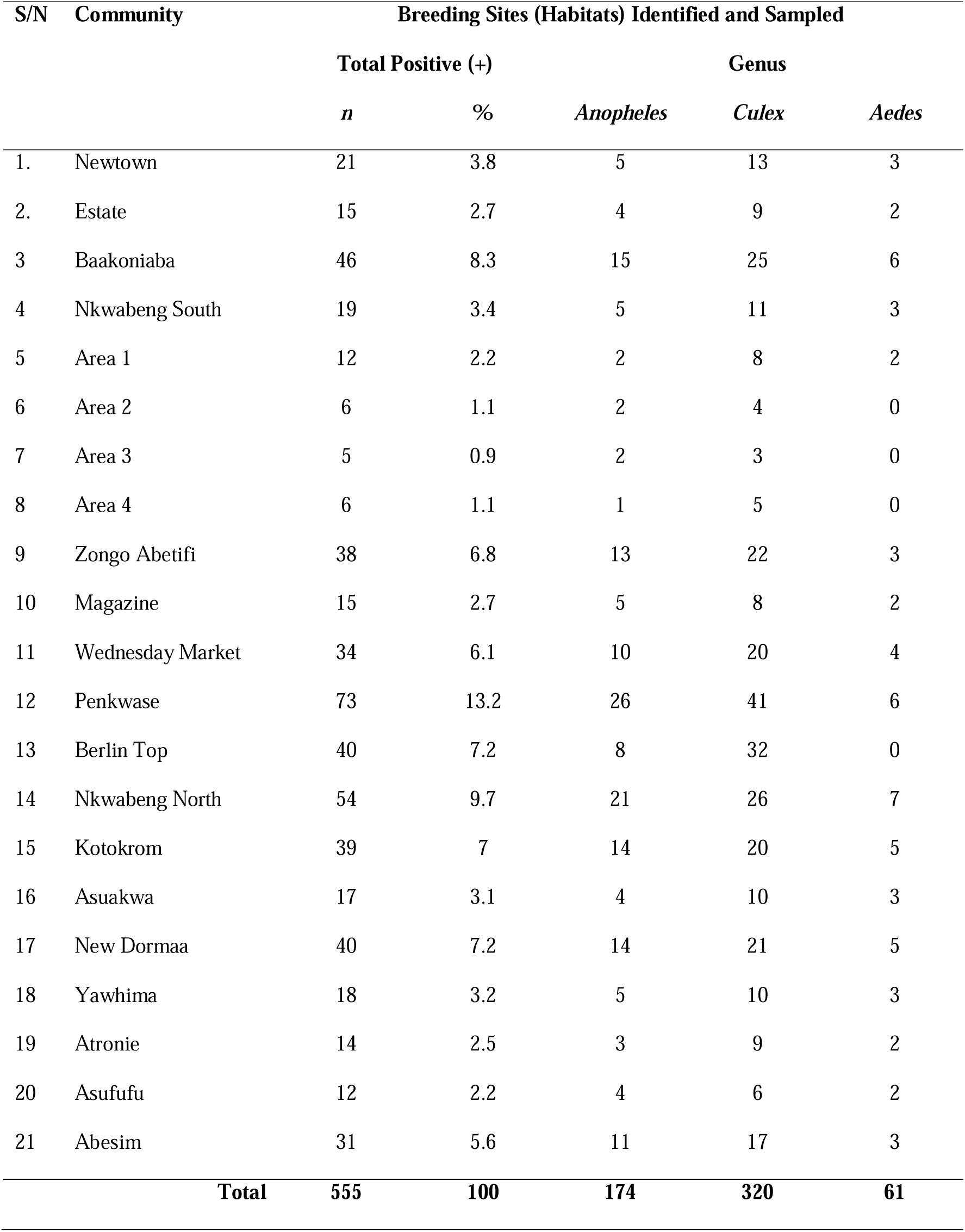
Distribution of Positive Mosquito Breeding Sites and Genus Composition Across Communities in the Sunyani Municipality.

| S/N | Community | Breeding Sites (Habitats) Identified and Sampled |  |  |  |  |
| --- | --- | --- | --- | --- | --- | --- |
|  |  | Total Positive (+) |  | Genus |  |  |
|  |  | *n* | % | *Anopheles* | *Culex* | *Aedes* |
| 1. | Newtown | 21 | 3.8 | 5 | 13 | 3 |
| 2. | Estate | 15 | 2.7 | 4 | 9 | 2 |
| 3 | Baakoniaba | 46 | 8.3 | 15 | 25 | 6 |
| 4 | Nkwabeng South | 19 | 3.4 | 5 | 11 | 3 |
| 5 | Area 1 | 12 | 2.2 | 2 | 8 | 2 |
| 6 | Area 2 | 6 | 1.1 | 2 | 4 | 0 |
| 7 | Area 3 | 5 | 0.9 | 2 | 3 | 0 |
| 8 | Area 4 | 6 | 1.1 | 1 | 5 | 0 |
| 9 | Zongo Abetifi | 38 | 6.8 | 13 | 22 | 3 |
| 10 | Magazine | 15 | 2.7 | 5 | 8 | 2 |
| 11 | Wednesday Market | 34 | 6.1 | 10 | 20 | 4 |
| 12 | Penkwase | 73 | 13.2 | 26 | 41 | 6 |
| 13 | Berlin Top | 40 | 7.2 | 8 | 32 | 0 |
| 14 | Nkwabeng North | 54 | 9.7 | 21 | 26 | 7 |
| 15 | Kotokrom | 39 | 7 | 14 | 20 | 5 |
| 16 | Asuakwa | 17 | 3.1 | 4 | 10 | 3 |
| 17 | New Dormaa | 40 | 7.2 | 14 | 21 | 5 |
| 18 | Yawhima | 18 | 3.2 | 5 | 10 | 3 |
| 19 | Atronie | 14 | 2.5 | 3 | 9 | 2 |
| 20 | Asufufu | 12 | 2.2 | 4 | 6 | 2 |
| 21 | Abesim | 31 | 5.6 | 11 | 17 | 3 |
| **Total** |  | **555** | **100** | **174** | **320** | **61** |

Further, the spatial data revealed areas with high concentrations of breeding sites, particularly in urbanized communities such as Penkwase and Nkwabeng North, which exhibited increased mosquito density. By integrating geospatial data and laboratory analysis, the mapping process identified patterns of habitat suitability and mosquito proliferation, facilitating targeted interventions. These findings are critical for optimizing vector control strategies and mitigating mosquito-borne disease transmission within the Municipality.

**Fig. 4** provides a detailed geospatial visualization of mosquito larval collection sites across Sunyani Municipality, emphasizing high-risk zones for mosquito breeding. The map overlays community locations, species composition, and spatial patterns to highlight areas with significant mosquito larval proliferation. Communities are marked with red dots, indicating the locations of larval collection sites, while pie charts within the map represent the genus composition (*Anopheles*, *Culex*, and *Aedes*) observed in each community. The Municipal boundary is delineated with a red line, distinguishing Sunyani from neighbouring districts.

**Fig. 4.**
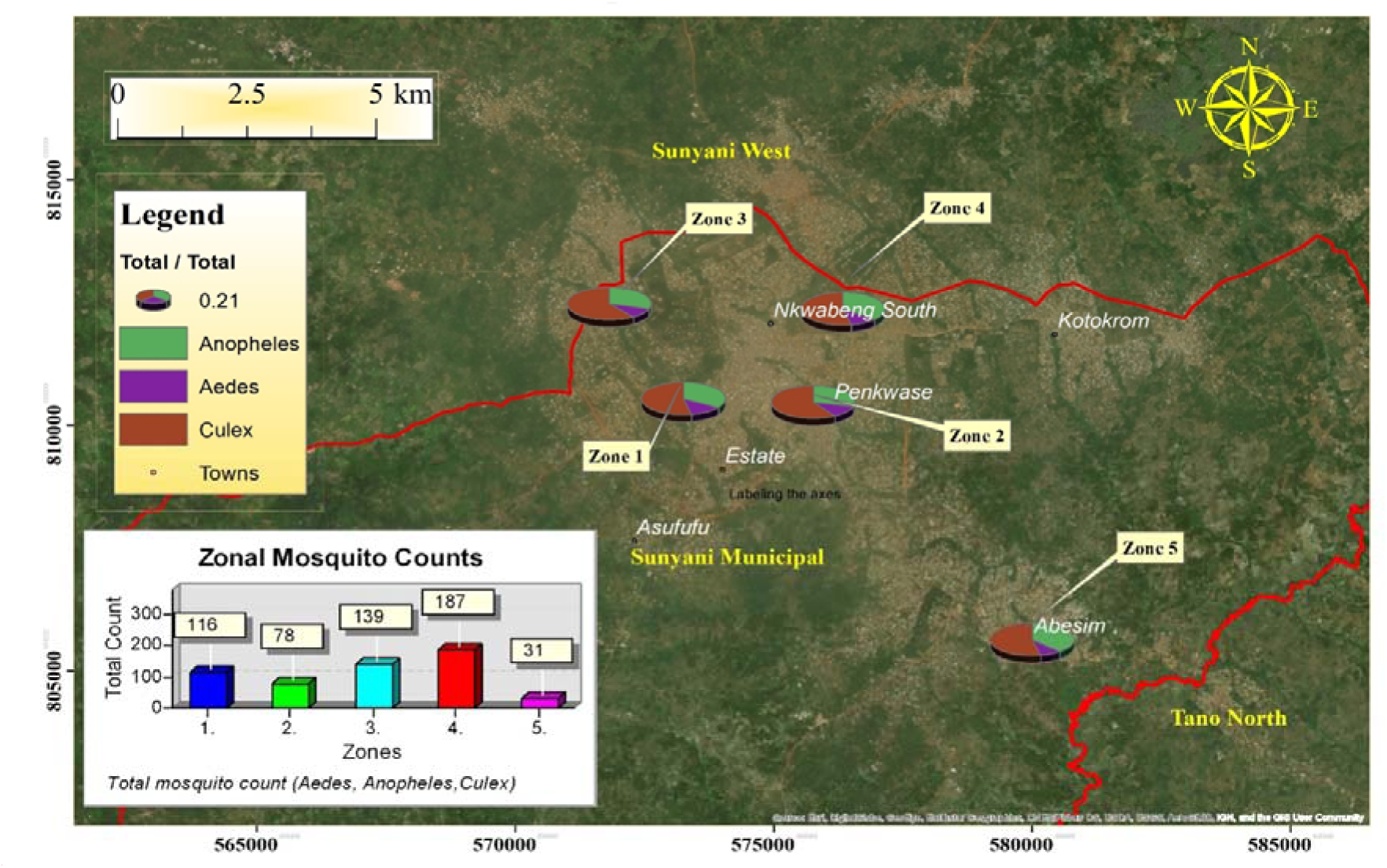
Geospatial distribution of mosquito larval collection sites across Sunyani Municipality.

Penkwase and Nkwabeng North emerge as prominent hotspots for mosquito larval activity, with these communities accounting for the highest density of positive breeding sites. Penkwase, in particular, recorded 73 positive sites, representing 13.2% of the total breeding sites sampled, attributed to its urban infrastructure and poorly maintained drainage systems that create stagnant water favorable for mosquito breeding. Nkwabeng North, with 54 positive sites (9.7%), similarly features environmental and anthropogenic conditions conducive to mosquito proliferation. The clustering of breeding sites in these areas underscores the impact of localized factors, such as improper waste management, high population density, and water stagnation, on mosquito habitat formation.

### Genus-Specific Preferences Across Mosquito Breeding Sites

The study reveal interconnected patterns in mosquito breeding site characteristics, species distribution, and larval abundance, underscoring the ecological and spatial dynamics of mosquito populations in the Sunyani Municipality. Among the 174 breeding sites positive for *Anopheles* larvae, the majority were associated with wetlands (64%), followed by natural drains with partially clean water (19%), and burst or leaking water distribution pipelines (17%) (**Fig. 5A**). These results highlight the preference of *Anopheles* mosquitoes for natural and semi-natural aquatic habitats, where stagnant water conditions are favorable for their larval development. This emphasizes the importance of targeted habitat management, especially in wetland areas, to reduce *Anopheles* populations effectively. When considering species distribution across all 555 positive breeding sites, *Culex* larvae were the most prevalent, found in 320 sites (58%), followed by *Anopheles* in 174 sites (31%), and *Aedes* in 61 sites (11%) (**Fig. 5B**).

**Fig. 5:**
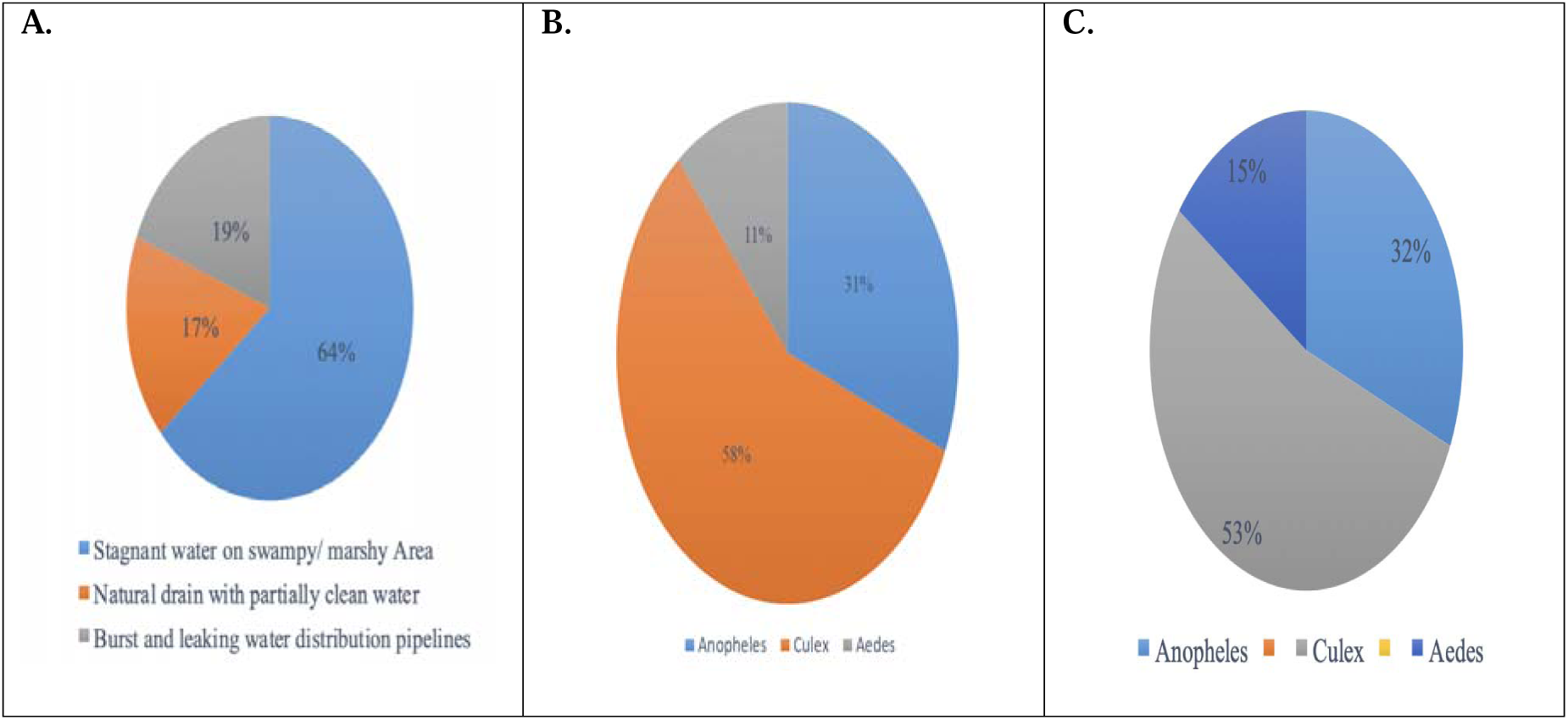
(A) Categories of mosquito breeding sites sampled positive for *Anopheles*, showing the distribution across wetlands, natural drains, and leaking water pipelines; (B) Positive breeding sites (habitats) identified for all mosquito genus, illustrating the proportion of sites supporting *Anopheles*, *Culex*, and *Aedes* larvae; and (C) Total mosquito larvae collected for all genus, highlighting the dominance of *Culex* larvae compared to *Anopheles* and *Aedes*.

The larval collection data further reinforces these patterns, with a total of 1,555 larvae sampled across the study area. Of these, *Culex* accounted for 830 (53%), *Anopheles* for 490 (32%), and *Aedes* for 235 (15%) (**Fig. 5C**). The predominance of *Culex* species is consistent with their ability to exploit a wide range of habitats, including polluted water sources. In contrast, *Anopheles* and *Aedes* larvae were confined to more specific ecological niches, such as clean natural water bodies and urban containers, respectively.

The larval densities (LD) was measured by the total number of larvae divided by the number of dips, which was 0.4.

### Assessment of Potential Mosquito Breeding Sites

A total of 819 potential mosquito breeding sites were identified as negative for larvae during the study period, representing habitats where no active larval presence was detected at the time of sampling. However, these sites were categorized based on their likelihood of becoming active mosquito breeding sites under favorable environmental conditions. Among these, 425 sites (52%) were identified as likely habitats for *Culex* mosquitoes, 238 sites (29%) for *Anopheles*, and 156 sites (19%) for *Aedes*.

The combination of field observations and geospatial mapping provided comprehensive data on mosquito breeding dynamics, laying the foundation for designing targeted and sustainable vector control strategies tailored to the unique ecological and anthropogenic conditions of the Sunyani Municipality.

## Discussion

The field survey identified significant mosquito breeding sites across the Sunyani Municipality, with a particular focus on *Anopheles* larvae. Out of the positive breeding sites sampled, 64% were located in wetlands (swampy/marshy areas), 19% in natural drains with partially clean water, and 17% in burst and leaking water distribution pipelines. These findings underscore the importance of natural and semi-natural aquatic habitats for *Anopheles* mosquito proliferation. The preference for wetlands as breeding sites reflects their ecological reliance on stagnant and relatively clean water, while the presence of larvae in natural drains and pipelines highlights the adaptability of *Anopheles* to semi-natural and anthropogenic habitats during the dry season **(Fig. 6A, B, D)**. In contrast, *Culex* mosquito larvae were identified in diverse habitats, including greywater in engineered drains, eroded natural drains, and artificial containers such as yellow gallons, discarded lorry tires, and metal barrels. The role of environmental mismanagement in creating mosquito-friendly conditions is evident from the presence of liquid waste, solid waste, and poor greywater management near these potential breeding sites **(Fig. 6C, D)**. Liquid waste, often associated with urban runoff and leaking pipes, was frequently observed near areas likely to support Culex mosquitoes. This findings corroborate to a study which pointed out that, the broad adaptability of *Culex* species to polluted environments emphasizes the role of poor sanitation and waste management in sustaining their populations ^[37]^. Similarly, *Aedes* mosquitoes were found in artificial habitats, including yellow gallons, discarded barrels, lorry tires and seats, empty cans, and greywater in drains **(Fig. 7)**. As container-breeding species, *Aedes aegypti* and *Aedes albopictus* are particularly adept at exploiting artificial habitats. Their ability to lay desiccation-resistant eggs on container walls enables them to survive and thrive in water-filled receptacles when conditions become favorable ^[37]^.

**Fig. 6:**
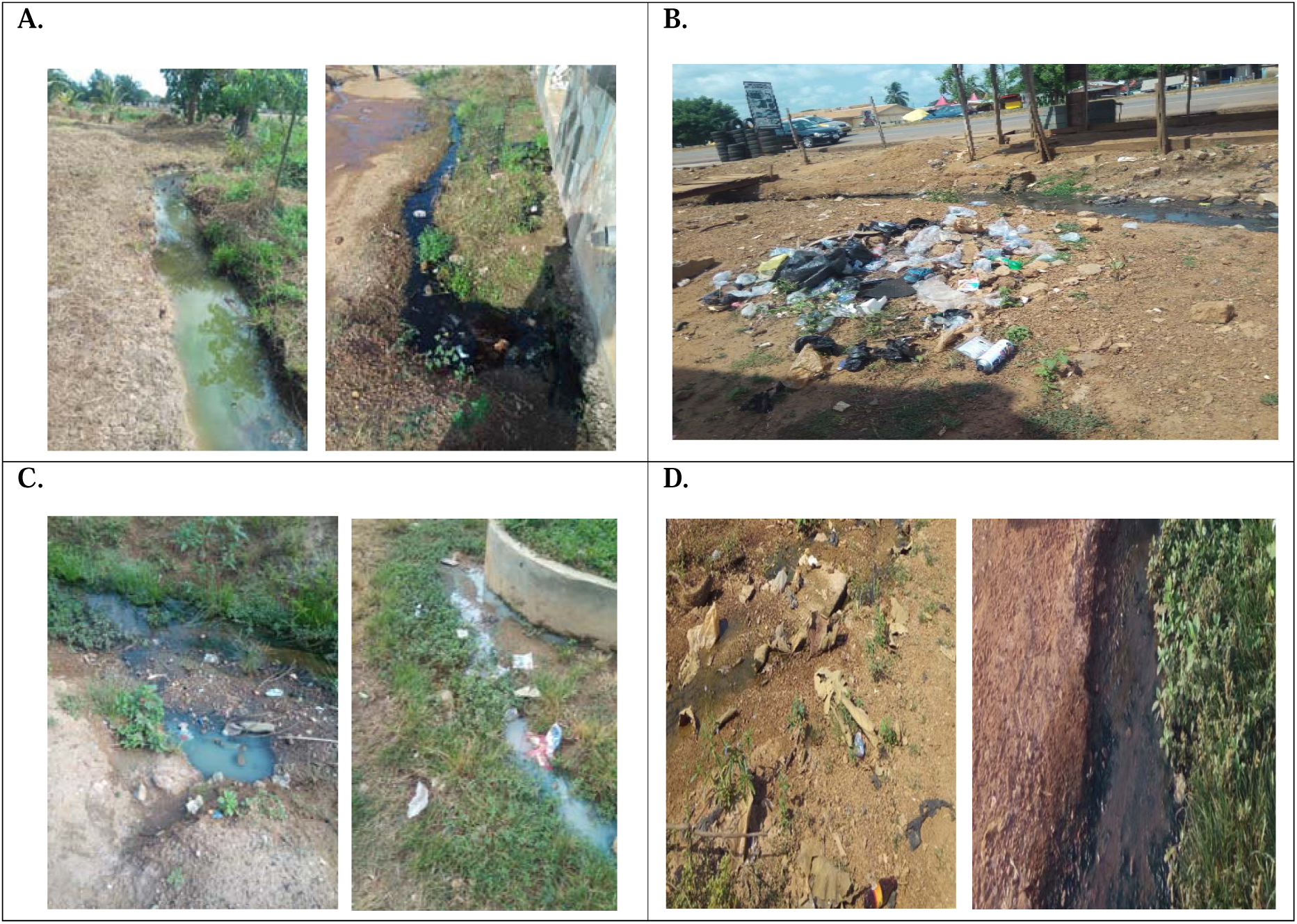
Environmental factors influencing mosquito breeding sites in Sunyani Municipality. (A) Liquid waste near breeding sites sampled positive for *Anopheles* and *Culex* mosquitoes; (B) Solid waste observed close to breeding sites positive for *Anopheles* and *Culex* mosquitoes; (C) Poor greywater management around breeding sites supporting *Culex* and *Aedes* mosquitoes; and (D) Littering observed near breeding sites positive for *Anopheles* and *Culex* mosquitoes. These environmental conditions highlight the role of mismanagement in facilitating mosquito habitat formation.

**Fig. 7:**
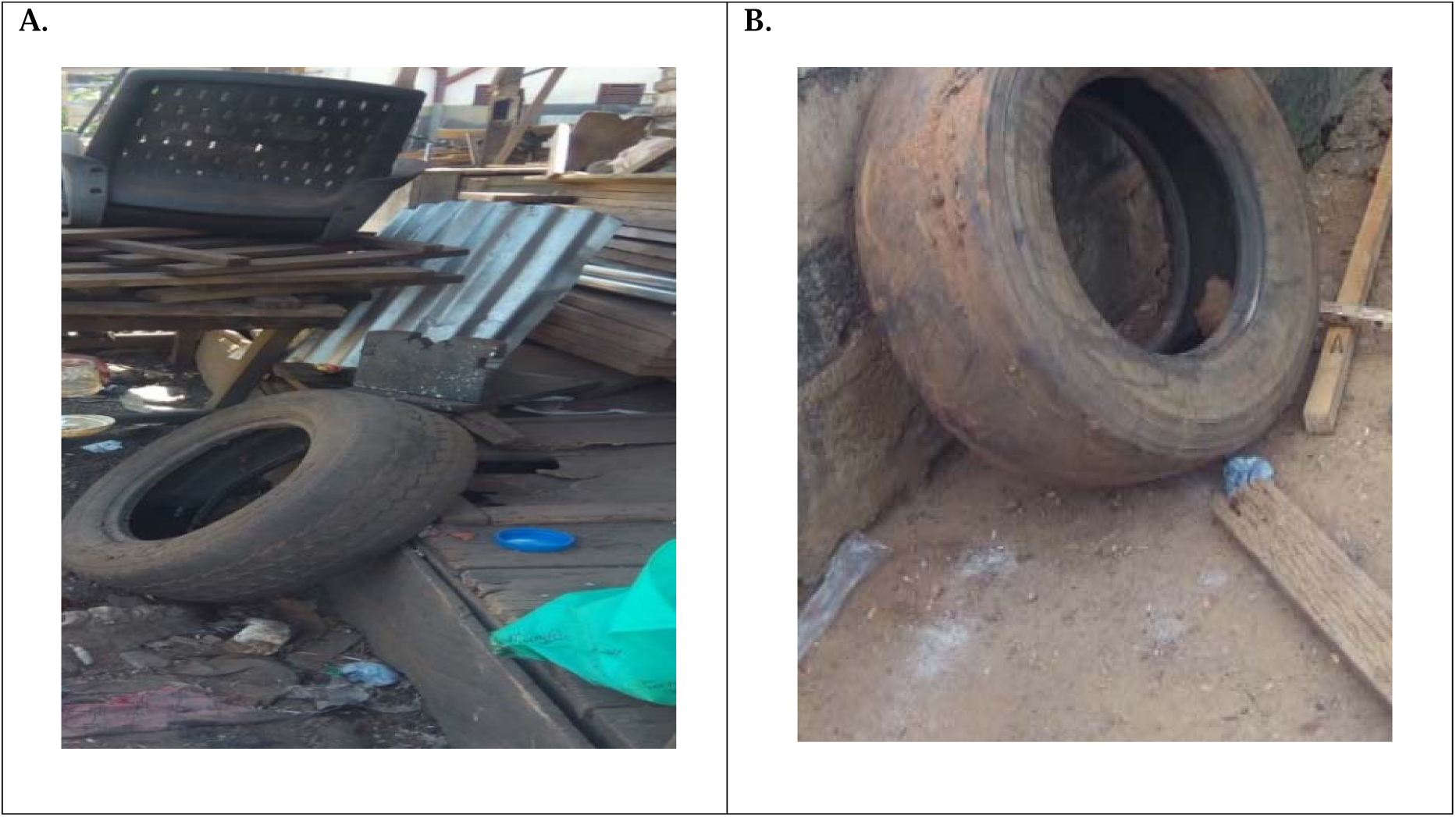
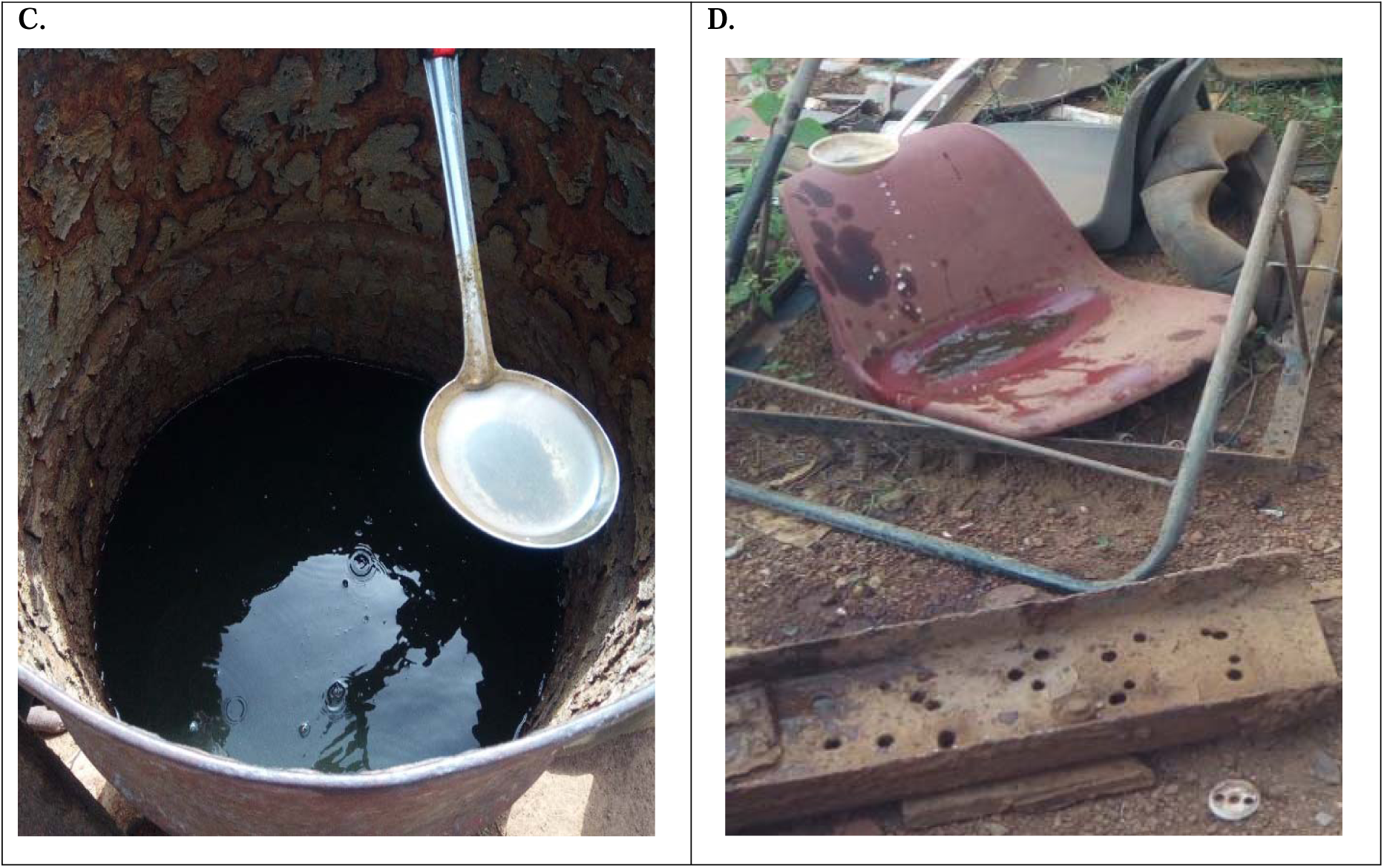
Environmental factors influencing mosquito breeding sites in Sunyani Municipality. (A) Accumulated water in a discarded lorry tire in a carpentry shop sampled positive for *Aedes* mosquitoes; (B) Accumulated water in a discarded lorry tire on a dwelling premises (house) sampled positive for *Aedes* mosquitoes; (C) Accumulated water in an open barrel on a dwelling premises (house) sampled positive for *Culex* and *Aedes* mosquitoes; and (D) stagnant water accumulated on a discarded car seat at a mechanic shop sampled positive for *Aedes* mosquitoes.

The timing of the survey, conducted during the dry season (December 2019 to February 2020), likely influenced the observed habitat preferences. Many temporary and shallow water bodies that *Anopheles* species typically exploit during the rainy season were absent, resulting in the concentration of larvae in semi-permanent habitats like wetlands and natural drains **(Fig. 6A, B, D)**. This seasonality highlights the critical importance of year-round surveillance and targeted vector control interventions to account for temporal changes in mosquito breeding sites. Mosquitoes, including *Anopheles*, undergo complex life cycles requiring aquatic habitats for egg, larval, and pupal development. Female *Anopheles* mosquitoes lay eggs in water, which hatch into larvae that feed and grow before pupating and emerging as adults. The reliance on aquatic habitats makes environmental management and sanitation critical components of vector control. Effective and timely interventions, including habitat modification, drainage improvement, and waste management, have proven successful in reducing mosquito breeding and eradicating known habitats ^[39]^. Larval source management (LSM), which targets the immature stages of mosquito development, has been documented as an efficient strategy for controlling *Anopheles* populations, particularly in natural wetlands ^[40]^. This approach, combined with modern technologies such as Geographic Positioning Systems (GPS) and Geographic Information Systems (GIS), enhances the mapping and profiling of mosquito breeding sites to inform targeted interventions ^[14]^. Such tools are instrumental in designing evidence-based, species-specific control strategies. The endemic nature of malaria in the study area is reinforced by the persistent presence of diverse *Anopheles* breeding sites. This emphasizes the need for robust vector control strategies and enforcement of public health regulations, such as Section 41 of the Public Health Act (PHA), 2012 (Act 851), which mandates property owners to prevent conditions favorable for mosquito breeding ^[41]^. These regulatory measures, when combined with community education and integrated vector management (IVM) programs, can significantly reduce mosquito populations and the associated disease burden in the Municipality.

Out of the 555 positive breeding sites identified and sampled in the Sunyani Municipality, a significant proportion was concentrated in specific communities. Penkwase recorded the highest percentage of positive sites (13.2%), followed by Nkwabeng North (9.7%), Baakoniaba (8.3%), and New Dormaa and Berlin Top (both at 7.2%). This predominance can be attributed to Penkwase’s dense population, urban infrastructure, and prevalence of poorly managed water storage and drainage systems, which create ideal conditions for mosquito breeding. Other communities, including Kotokrom (7%), Zongo Abetifi (6.8%), and Wednesday Market (6.1%), also contributed considerably to the total positive breeding sites. These communities are characterized by the presence of extensive natural wetlands (swampy/marshy areas), which provide ideal conditions for mosquito breeding, particularly for *Anopheles* species. In contrast, communities such as Area One, Area Two, Area Four, and Area Three recorded the lowest proportions, ranging from 2.2% to 0.9%. These areas are predominantly located in the Central Business District (CBD) and are devoid of natural wetlands, with positive samples predominantly originating from artificial habitats such as burst and leaking water pipelines and partially clean natural or eroded drains. The high prevalence of breeding sites in communities with natural wetlands underscores the role of these habitats in sustaining mosquito populations. However, the lower prevalence may be due to better water management practices, fewer anthropogenic water recipients, or environmental factors less conducive to mosquito breeding. The disparities in the number of positive sites among communities suggest that localized factors, including population density, sanitation practices, and the availability of stagnant water, play a critical role in influencing mosquito breeding site availability. This finding aligns with studies suggesting that stagnant water in wetlands, rice fields, and other natural water bodies serves as a principal breeding site for mosquito vectors, particularly *Anopheles* species ^[42]^. Other breeding sites mentioned in recent literature include water accumulation on leaves, uncovered water receptacles, and polluted water, reflecting a broader range of potential habitats that require attention in vector control programs ^[42].^ The dominance of wetlands in communities such as Penkwase and Nkwabeng North suggests poor environmental management and sanitation, which exacerbate mosquito breeding. These findings point to the urgent need for targeted environmental interventions to reduce mosquito breeding sites, including improving drainage systems, eliminating stagnant water, and educating the community on best practices for waste and water management.

The presence of female *Anopheles* mosquitoes, the primary malaria vectors in Sub-Saharan Africa, is of particular concern ^[44–45]^. Among these, *Anopheles gambiae* is the most efficient malaria vector due to its anthropophagic and endophilic behaviours, which enhance its ability to transmit *Plasmodium falciparum* effectively ^[46–47].^ Its adaptability to both natural and artificial habitats further underscore the importance of robust mosquito surveillance, including morphological identification, to support species-specific interventions. Mapping and profiling mosquito breeding sites, coupled with morphological identification of species involved in malaria transmission, are essential for designing and implementing effective vector control strategies. These approaches not only inform the ecological and biological characteristics of mosquito populations but also provide critical insights for tailoring Social and Behavioural Change Communication (SBCC) messages aimed at educating local communities on malaria control and prevention. Furthermore, active surveillance of vector control interventions, such as larvicides and insecticides, ensures that environmental impacts are minimized while maximizing the effectiveness of Integrated Pest Management (IPM) programs ^[43]^. The high prevalence of mosquito breeding sites in certain communities reflects deficiencies in environmental management, highlighting the need for community-level interventions to reduce mosquito populations and mitigate the risk of vector-borne diseases (VBDs). These interventions, when integrated with effective surveillance and community education, have the potential to significantly reduce the burden of malaria and other mosquito-borne diseases in Sunyani Municipality.

The predominance of *Culex* mosquitoes reflects the adaptability of this species to polluted and artificial habitats, exacerbated by poor waste management practices. The abundance of *Culex* larvae can be attributed to the widespread availability of solid and liquid waste, such as discarded containers, plastic materials, and stagnant water in drains, which serve as ideal breeding grounds **(Fig. 6 and 7C)**. These findings are consistent with the study by Agyemang-Badu et al ^[42]^, which highlighted the role of poor sanitation, including the presence of empty gallons, cans, and plastic containers, in creating mosquito-friendly environments. Addressing these waste management deficiencies is essential to reducing *Culex* populations and the associated risk of diseases such as lymphatic filariasis. The presence of *Anopheles* larvae, comprising 32% of the total collection, underscores the continued risk of malaria transmission in the study area. Female *Anopheles* mosquitoes, the primary malaria vectors in Africa, exhibit seasonal abundance, with the former thriving during the wet season and the latter during the dry season ^[53–54]^. These species are highly adapted to tropical climates with suitable temperature and rainfall patterns, facilitating malaria transmission across West and Central Africa. The ability of female *Anopheles* mosquitoes to exploit natural and semi-natural aquatic habitats, coupled with their anthropophagic behaviour, makes them particularly efficient vectors of *Plasmodium falciparum*. Understanding these ecological and behavioural traits is critical for implementing targeted larval source management and other vector control strategies.

The 15% contribution of *Aedes* larvae to the total mosquito population is significant, given the role of *Aedes* species, particularly *Ae. albopictus*, in transmitting arboviral diseases such as dengue, chikunguya, and Zika virus. The rapid population growth of *Aedes* mosquitoes, driven by extreme temperature conditions and other effects of climate change, poses a growing public health threat ^[51]^. Changes in ecological conditions associated with climate change, such as altered rainfall patterns and increased temperatures, may expand the geographic range and breeding potential of *Aedes* mosquitoes, thereby increasing the risk of disease outbreaks. This highlights the need for proactive measures to monitor and control *Aedes* populations, particularly in urban and peri-urban areas where artificial containers provide abundant breeding sites. Effective mosquito control requires an in-depth understanding of species-specific breeding habitats and ecological requirements. As noted by ^[52]^, targeting the appropriate breeding habitats is essential for successful intervention at either the larval or adult stages. Given the findings of this study, control measures should prioritize the improvement of waste management systems to reduce *Culex* breeding habitats, alongside habitat modification and larval source management to address *Anopheles* populations. For *Aedes*, community engagement and public health education are crucial for reducing artificial container breeding sites. By addressing these challenges, integrated vector management programs can significantly mitigate the risk of mosquito-borne diseases in the study area.

This dominance of *Culex* mosquitoes can be attributed to their adaptability to polluted and anthropogenic habitats, such as poorly managed drains and waterlogged urban areas. *Aedes* larvae, on the other hand, were predominantly associated with artificial containers and urban environments, reflecting their ecological niche preference.

The predominance of potential *Culex* breeding sites reflects their ecological preference for polluted and stagnant water bodies, such as dirty and choked gutters, which were more pronounced during the dry season (December 2019 to February 2020). This finding aligns with Service, ^[27]^, which notes that *Culex* mosquitoes breed in polluted drains, septic tanks, and choked water systems, emphasizing the role of poor waste management and sanitation in their proliferation. The potential breeding sites for *Anopheles* mosquitoes were associated with clean and stagnant water bodies, including shallow and fresh water devoid of shade, as well as temporary water sources like potholes, rice fields, excavations, catch pits, vegetable farms, ponds, and tidal swamps. These findings reflect the ecological characteristics of *Anopheles* mosquitoes, which thrive in relatively clean aquatic habitats. However, the absence of active larvae in these sites during the dry season highlights the seasonal dependence of *Anopheles* breeding, with their habitats more abundant during the rainy season. The role of settlement patterns near wetlands and swampy areas in contributing to *Anopheles* breeding was emphasized by participants in a related study, who noted that proximity to such areas significantly increases the risk of malaria and other mosquito-borne diseases ^[42]^.

In this study, *Aedes* mosquito breeding sites were primarily associated with container habitats such as discarded barrels, lorry tires and seats, empty cans, and greywater in drains. Their preference for artificial containers and clean stagnant water underscores the importance of addressing urban waste and household water management practices to reduce the proliferation of *Aedes* mosquitoes **(Fig. 7)**. These findings corroborate earlier research identifying the role of poorly managed water storage containers and discarded materials in facilitating *Aedes* breeding ^[27]^. The seasonal timing of the larval sampling influenced the identification of potential breeding sites, as dry season conditions often reduce the availability of natural and temporary water bodies, while anthropogenic habitats like dirty gutters and containers remain active. This seasonal variability highlights the need for year-round monitoring to capture the full spectrum of mosquito breeding habitats. The findings reinforce the importance of targeted interventions based on species-specific habitat preferences. For *Culex*, improving drainage systems and addressing solid waste management are critical. For *Anopheles*, interventions should focus on modifying wetland areas and managing temporary water sources, particularly during the rainy season. For *Aedes*, reducing artificial container habitats through community awareness campaigns and regular waste disposal is essential. These tailored strategies, integrated with community engagement and public health education, are vital for reducing the risk of mosquito-borne diseases in the Municipality.

However, the researchers could not identified the laboratory reared adult mosquitoes to the species level due to factors beyond their SBCC control such as financial constraints of securing rDNA Polymerase Chain Reaction (PCR) machine and local lockdown and travel restricts to access well-resourced research laboratories in Ghana at the time of the study, which sets as a key limitation of the study.

## Conclusion

This study highlights the critical role of specific habitats, including wetlands (swampy/marshy areas), natural drains with partially clean water, and burst or leaking water distribution pipelines, in sustaining *Anopheles* mosquito populations in the Sunyani Municipality. The predominance of *Culex* in anthropogenic habitats, such as greywater drains and discarded containers, underscores the influence of poor sanitation and waste management practices in promoting mosquito proliferation. The study also confirms that stagnant and accumulated water remains a critical factor driving mosquito breeding and survival, thereby sustaining vector populations and disease transmission. To mitigate mosquito breeding and reduce vector-borne disease prevalence, integrated strategies are essential. Regulatory authorities, including the Physical Planning Department (PPD) of the Sunyani Municipal Assembly (SMA), must enforce the Land and Spatial Planning Act (LSPA) and Building Regulations (BL) to restrict construction near natural wetlands. Concurrently, strict enforcement of Environmental Sanitation By-Laws (ESBL) by licensed Environmental Health Officers (EHOs) is crucial to eliminate potential breeding habitats. Public health interventions should also include community awareness and education campaigns to promote proper waste disposal and water management practices. These findings provide valuable insights into the environmental and anthropogenic factors influencing mosquito breeding, offering a foundation for targeted interventions and policy making. By integrating habitat management, regulatory enforcement, and community engagement, sustainable reductions in mosquito populations and associated diseases can be achieved in the Sunyani Municipality.

## Abbreviations

ACE: African Centre of Excellence
BL: Building Regulations
CBD: Central Business District
CHRPE: Committee on Human Research Publication and Ethics
EHOs: Environmental Health Officers
ESBL: Environmental Sanitation By-Laws
GIS: Geographic Information System
GPS: Geographic Positioning Systems
IPM: Integrated Pest Management
IVM: Integrated Vector Management
KNUST: Kwame Nkrumah University of Science and Technology
LSPA: Land and Spatial Planning Act
LD: Larval Densities
MBDs: Mosquito-Borne Diseases
PPD: Physical Planning Department
PCR: Polymerase Chain Reaction
PHA: Public Health Act
RWESCK: Regional Water and Environmental Sanitation Centre-Kumasi
SBCC: Social and Behavioural Change Communication
SMA: Sunyani Municipal Assembly
TSMA: Tano South Municipal Assembly
VBDs: Vector-Borne Diseases
WB: World Bank

## Acknowledgments

The authors would like to thank all the data collection assistants who wholeheartedly participated in the field data collection across the Sunyani Municipality. Also, much thanks goes to Ms. Martha Okrah and Mr. Joseph Kwarko-Kyei for their immense contribution during the data collection and processing. Further thanks goes to All the Professors, Senior Lecturers and Staff of Regional Water and Environmental Sanitation Centre-Kumasi (RWESCK), Kwame Nkrumah University of Science and Technology (KNUST), Kumasi for their massive contribution and support to the success of this project.

## Obligatory funder statement for research article publication

‘’This study was funded by the Regional Water and Environmental Sanitation Centre Kumasi (RWESCK) at the Kwame Nkrumah University of Science and Technology (KNUST), Kumasi with funding from the Ghana Government through the World Bank (WB) under the Africa Centres of Excellence project.’ The views expressed in this study do not reflect those of the World Bank, Ghana Government, and RWESCK KNUST.’’

## Data availability statement

The datasets generated and/or analysed during the current study are available from the corresponding author upon reasonable request.

## Author contributions

Conceptualization, SYAB, EA, and SOK; methodology, SYAB, EA, SOK and NCD; validation EA, SOK and JYWD; formal analysis, SYAB, NCD and JYWD; project/data collection, SYAB and SO; data curation, EA, SOK and JYWD; writing original draft preparation, SYAB, JYWD and SO; writing review and editing, EA, SOK and NCD; supervision, EA and SOK; funding acquisition, SYAB, EA and SOK. All authors have read and agreed to the published version of the manuscript.

## Informed consent statement

Not applicable.

## Consent for publication

Not applicable.

## Supplementary materials

Not applicable.

